# LDL-apheresis as an alternate method for plasma LPS purification in healthy volunteers, dyslipidemic and septic patients

**DOI:** 10.1101/2020.10.07.20206771

**Authors:** Auguste Dargent, Jean-Paul Pais de Barros, Samir Saheb, Randa Bittar, Wilfried Le Goff, Alain Carrié, Thomas Gautier, Isabelle Fournel, Anne Laure Rerole, Hélène Choubley, David Masson, Laurent Lagrost, Jean-Pierre Quenot

## Abstract

Lipopolysaccharide (LPS) is a key player for innate immunity activation. It is therefore a prime target for sepsis treatment, as antibiotics are not sufficient to improve outcome during septic shock. Extracorporeal removal method by polymyxin B hemoperfusion (PMX-DHP) is used in Japan, but recent trials failed to show a significant lowering of circulating LPS levels after PMX-DHP therapy. PMX-DHP has a direct effect on LPS molecules. However, LPS is not present in a free form in the circulation, as it is mainly carried by lipoproteins, including low density lipoproteins (LDL). Lipoproteins are critical for physiological LPS clearance, as LPS are carried by low density lipoproteins (LDL) to the liver for elimination. We hypothesized that LDL-apheresis can be an alternate method for LPS removal. We demonstrated first *in vitro* that LDL apheresis microbeads are almost as efficient as PMX beads to reduce LPS concentration in LPS-spiked human plasma, whereas it is not active in phosphate-buffered saline. We found that PMX was also adsorbing lipoproteins, although less specifically. Then, we found that endogenous LPS of patients treated by LDL-apheresis for familial hypercholesterolemia is also removed during their LDL-apheresis sessions, both with electrostatic-based devices and filtration devices. Finally, LPS circulating in the plasma of septic shock and severe sepsis patients with gram-negative bacteremia was also removed *in vitro* by LDL adsorption. Overall, these results underline the importance of lipoproteins for LPS clearance, making them a prime target to study and treat endotoxemia-related conditions.

## Introduction

Lipopolysaccharide (LPS), a component of the outer membrane of Gram-negative bacteria (GNB), is the most widely studied and one of the most potent pathogen-associated molecular patterns (PAMPs) (1). LPS is a potent trigger of the innate immune response during GNB sepsis, although its role was also demonstrated in non-GNB sepsis and non-septic conditions (1, 2), probably because of endogenous sources of LPS magnified by shock, such as bacterial translocation from the gut. LPS has been considered as a therapeutic target for sepsis ever since it was recognized (3). Monoclonal anti-LPS antibodies failed in most clinical studies, partly because of poor specificity (4). Thereafter, encouraging results were obtained with extracorporeal removal by polymyxin (PMX) B hemoperfusion, developed in Japan (5). Unfortunately, further clinical trials did not confirm the efficacy of this treatment to improve clinical outcomes (6). Interestingly, the results of a recent, well-conducted clinical trial showed that circulating LPS levels were not significantly decreased after PMX hemoperfusion (7). In fact, even though the affinity of PMX for the lipid A moiety of LPS is strong in water, it is dramatically decreased when LPS is spiked in a protein solution, and even more in plasma (8).

Lipoproteins are well-known to play a major protective role during sepsis and inflammation (9). Due to its amphipathic properties, LPS is rapidly and massively (more than 75%) bound by plasma lipoproteins (10), partly explaining the low extraction capacity of polymyxin in plasma. In the same way, lipoproteins can also carry other important lipidic PAMPs such as lipoteichoic acid (LTA), a component of gram-positive bacteria cell wall (11). Lipoproteins enhance the clearance of LPS from plasma during sepsis (12): they bind, inactivate (13) and transport LPS molecules to the liver for biliary elimination (14). In rodents, sepsis is classically accompanied by a global cytokine-mediated increase of lipoproteins (15), whereas in humans, LDL and HDL cholesterol are decreased (16). Lipoprotein levels at the onset of sepsis affect the outcome of patients: a low HDL (high-density lipoprotein) level was recently found to be the best prognostic marker for adverse outcomes in a sepsis cohort (17). This corroborates earlier findings by our group showing that low cholesterol levels (both HDL and LDL) is a risk factor of sepsis and poor clinical outcome in patients undergoing cardiac surgery (18, 19). Indeed, LPS is first bound to HDL particles in the circulation, but its elimination requires first a transfer to LDL (low density lipoproteins) mediated by proteins such as LPS binding protein (LBP) or PLTP (phospholipid transfer protein)-mediated (14), and secondly internalization and degradation by hepatocytes via LDL receptor (LDLR-r) (20). We have termed this pathway reverse LPS transport (21) by analogy with cholesterol, and it may bear a critical role during human sepsis. We showed that PLTP-deficient mice had higher sepsis-induced mortality (22) and that recombinant PLTP could restore the biliary excretion capacity and improve survival of the animals (12). The role of LDL-r is showed in human sepsis where patients presenting loss-of-function mutations of the inhibitory protein PCSK9 (Proprotein convertase subtilisin/kexin 9) show increased survival (23) and increased LPS clearance (20), although PCSK9 inhibition in mice failed to reduce LPS-induced mortality (24). PCSK9-induced LDL-r decrease (and therefore decreased hepatic LDL uptake) is a feature shared by patients presenting sepsis-induced organ failure (25) and familial hypercholesterolemia (FH) (26). Patients with severe homozygous FH are successfully treated with extracorporeal LDL apheresis with columns using electrostatic adsorption of LDL’s apolipoprotein B (27). In the present study, we hypothesized that LDL adsorption can be used to remove LPS from plasma.

## Methods

### Healthy volunteers and critically ill patients

We used plasma samples from a biologic collection of a critically ill patients’ cohort from our medical Intensive Care Unit (ICU) in Dijon University hospital (IVOIRE study) (28). Patients (or their next of kin) were informed and signed an additional, specific consent form authorizing the conservation and utilization of biological samples. Plasma samples were collected on the day of inclusion (< 24 hours since ICU admission. The samples were processed following a strict protocol, 1 hour maximum after sampling, with protocolized centrifugation and immediate freezing at −80°C, then transferred for storage at the biobank facility of Dijon University Hospital (Centre de Ressources Biologiques Ferdinand Cabanne). Healthy ambulatory volunteers (n=49) were also recruited in this study to serve as controls. Each volunteer provided informed consent. Exclusion criteria for volunteers were: recent surgery (<30 days) or dental care (< 72 hours), any current or recent (<30 days) infectious episode with antibiotic therapy, current or recent (<30 days) immunosuppressant therapy including corticosteroids.

### Study approval

The institutional review board (Comité de Protection des Personnes Est I, Dijon) approved the protocol for the collection of LDL apheresis patients’ samples, and considered it to constitute routine clinical practice as no additional samples were drawn. The need for informed consent was waived, but all patients or their relatives were given clear information about the study, and their non-opposition was obtained. Collection of nominative data was approved by the national authority for the protection of privacy and personal data. Plasma samples from septic patients and healthy control subjects originate from a clinical study which received approval from the local Ethics Committee (Comité de Protection des Personnes Est I) under the number 2013/15 (ClinicalTrials.gov identifier NCT01907581). All patients and/or their next of kin were informed and consent was documented in the patients’ medical records by the investigator.

### LDL apheresis samples

Patients were recruited in a national reference center for LDL apheresis in Paris, France. Patients received information of the study goals and gave consent. No additional blood samples were drawn, as the remaining volume of plasma from the routine sample was used for the presented analyses. No further clinical data (other than epidemiologic) were collected.

### In vitro microbeads experiments

Individual plasma samples were spiked at a concentration of 500 ng/mL of O55:B5 E. coli LPS (Thermofisher Scientific, USA), and incubated for 2 hours at 37°C without agitation. Sampling for measurement of LPS, lipid profile and proteins at time 0 (T0) was made after incubation. Each plasma was then added to three different types of microbeads prepared in separated 2ml Eppendorf tubes. Microbeads used were: polymyxin B crosslinked 6% agarose (Separopore® 6B-CL) beads (Endotoxin AffisorbentTM, bioWorld, USA), DALI® system polyacrylate microspheres (Fresenius, Germany), and Separopore® 6B-CL (bioWorld, USA) as control. Microbeads were washed the same day with sterile saline (500ml for 10 grams) and stored at +4°C. In each Eppendorf tube, 400µl of plasma was added to 200mg of microbeads (1200µL of plasma per individual sample), following the plasma/substrate ratio used in clinical LDL-apheresis sessions 2. Eppendorf tubes were then incubated at 37°C for 2 hours with rotative agitation (25 RPM). Supernatant plasma was sampled after settling of the beads.

### Isolation of lipoprotein-deficient plasma fraction

Lipoprotein deficient plasma was used as an additional control matrix. Ten individual human plasma samples were used. Plasma density was adjusted to 1,21g/ml using solid KBr. Lipoprotein deficient plasmas were obtained by ultracentrifugation for 3 hours at 4°C, 100,000 rpm using a TLA-120.2 fixed-angle rotor in an optima TLX ultracentrifuge (Beckman Coulter). Infranatant was collected and dialyzed overnight against PBS buffer at 4°C.

### LPS analyses

LPS was quantified using a mass spectrometry (LC-MS/MS) patented method (EndoQuant) for detection of 3HM. For this purpose, two aliquots of plasma (50µl) are spiked with 4 pmol of internal standard (3-hydroxytridecanoic acid). For total 3HM quantitation, samples were hydrolysed with HCl 8 M for 3 h at 90°C. Free fatty acids were then extracted with hexane/ethyl acetate 3/2 mix, and resuspended in ethanol after vacuum evaporation. Fatty acid separation was performed in an Infinity 1290 HPLC binary system (Agilent) equipped with a ZORBAX SB-C18 C18 50 x 2.1 mm 1.8 µm column (Agilent) set at 30°C. MS/MS detection was performed using a QQQ 6490 triple quadruple mass spectrometer (Agilent) equipped with a Jet-Stream ESI source. Quantitation of 3HM was performed by negative SRM mode as previously described (29). The limulus amoebocytes lysate (LAL) assay was also used in parallel (LAL chromogenic endpoint assay, Hycult Biotech).

### Other analyses

Plasma total proteins, total cholesterol, LDL cholesterol, HDL cholesterol and triglycerides were analysed using an INDIKO® analyser (ThermoScientific, France).

### Statistical analysis

Data analysis were performed using GraphPad Prism 7. Normally distributed data were analyzed using paired, 2-tailed Student’s t test. Quantitative variables are presented as mean ± standard deviation (SD). All analyses were performed using with a significance threshold set at 0.05.

## Results

### LPS adsorption by LDL-apheresis microbeads in LPS-spiked healthy volunteers’ plasma

To determine if LDL apheresis columns can remove LPS from plasma, we first used plasma samples from 49 healthy volunteers, spiked and incubated for 2 hours at 37°C with 500ng/ml of O55:B5 E. coli LPS. Each individual LPS-spiked plasma was then incubated for 2 hours with agitation at 37°C after adding one of 3 different substrates: polymyxin B crosslinked 6% agarose (Separopore® 6B-CL) beads (Endotoxin Affisorbent™, bioWorld, USA), DALI® system (clinical LDL-apheresis column) polyacrylate microspheres (Fresenius, Germany), and Separopore® 6B-CL (bioWorld, USA) used as control. Control beads were used to normalize for the sample dilution induced by the hydrated substrates (sterile saline was used for priming of all substrates). For this reason, control substrate rather than baseline was used for reference in comparisons. Measurements of LPS were made using 2 different techniques. LAL remains the standard method for measuring endotoxin activity. However, LAL relevance in plasma is questioned (30), as there is interference with inhibitors, mainly lipoproteins as detailed earlier. We developed a mass spectrometry method retaining LAL’s sensitivity in plasma for LPS quantitation, using a specific compound, 3-hydroxy myristate (3HM). When LPS is spiked in plasma, 3HM concentration remains stable, whereas LAL reactivity is decreasing rapidly over time (29). Thus, 3HM concentration is probably a better indicator of the total amount of LPS in plasma (i.e. including lipoprotein-bound LPS), whereas LAL requires the lipid A moiety of LPS to be exposed. Endotoxin activity and 3HM levels both decreased significantly after incubation with DALI® beads compared to control beads, as well as after incubation with PMX (44.6% and 33.5% of control endotoxin activity, respectively, Figure 1A; and 72.8 ±11% and 61 ±13% of control 3HM levels, respectively, Figure 1C). A basic sensitivity analysis was performed: samples with high baseline levels of 3HM (above mean +SD) were removed (to eliminate a spurious effect caused by artificially high levels of LPS in the sample). The results remained unchanged with a similar, significant decrease of 3HM with DALI® and PMX beads compared to control (73.2 ±11% and 59.8 ±13%, respectively). The experiment was repeated in LPS-spiked phosphate buffered saline (PBS), and showed no effect of incubation with DALI beads, whereas incubation with PMX decreased LPS levels (significantly for 3HM, Figure 1B, non-significantly for endotoxin activity, Figure 1D). Additionally, DALI and PMX were also incubated with LPS-spiked lipoprotein-deficient plasma from 10 control plasma samples (Figure 2). After incubation with PMX, 3HM levels decrease was enhanced in lipoprotein-deficient plasma, as compared to whole plasma (58% reduction versus 40%, respectively). Conversely, efficiency of DALI microbeads was considerably reduced in lipoprotein-deficient plasma (11% reduction versus 27% in whole plasma), although 3HM decrease remained significant in this matrix (Figure 2).

**Figure 1:**
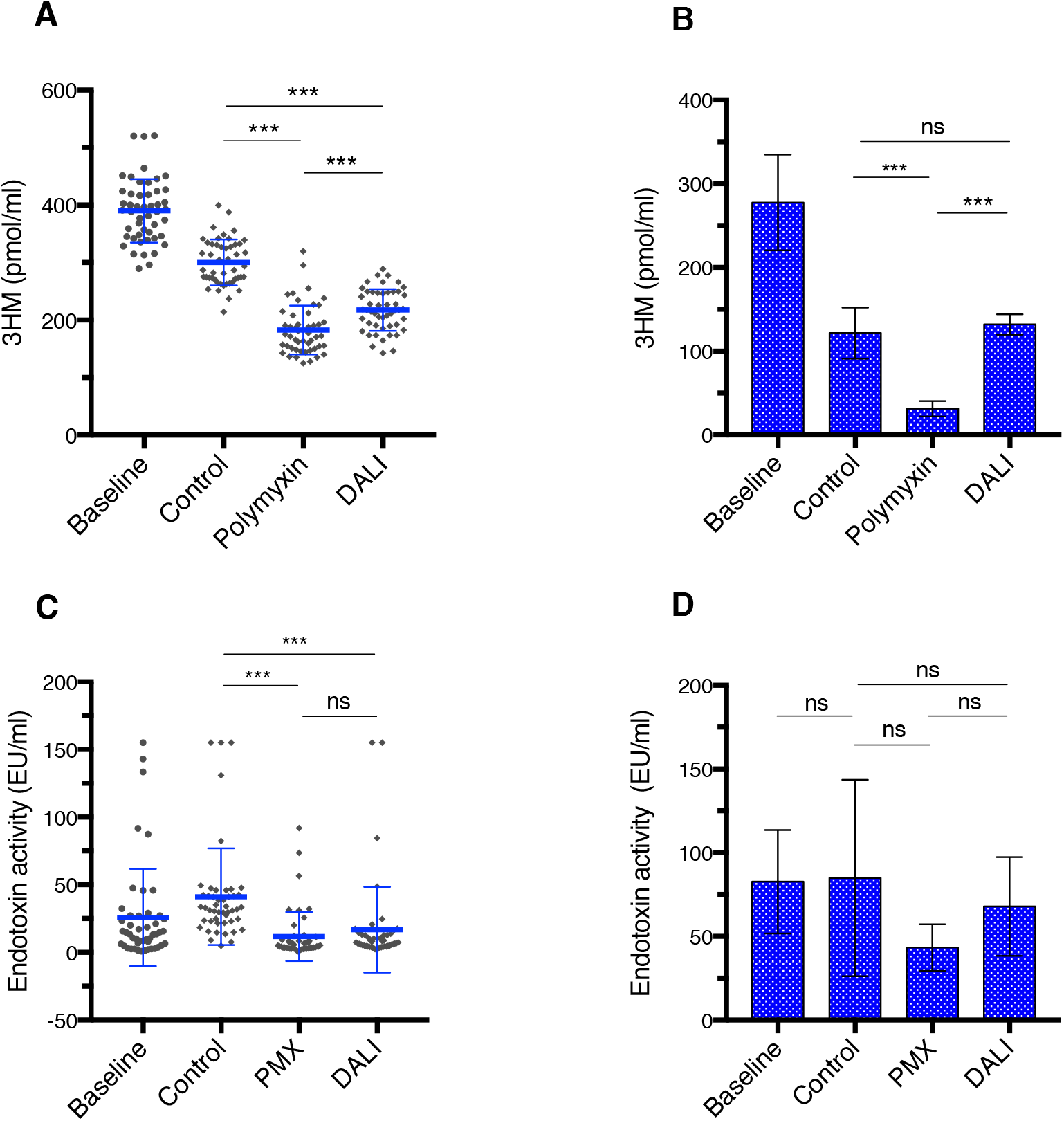
LDL apheresis microbeads are efficiently reducing LPS levels in spiked human plasma, but not PBS. LPS levels are decreasing after incubation with both DALI® and polymyxin (PMX) microbeads in LPS-spiked human plasma from 49 healthy volunteers, as assessed by 3HM levels (A), and LAL (limulus amoebocyte assay) (C). In spiked PBS (n=3), only incubation with PMX is reducing 3HM levels (B). LAL decrease with PMX did not reach significance in PBS (D). Data represent mean ± SD. *** p<.001 and ns: p>0.05 by two-tailed paired t-test.

**Figure 2:**
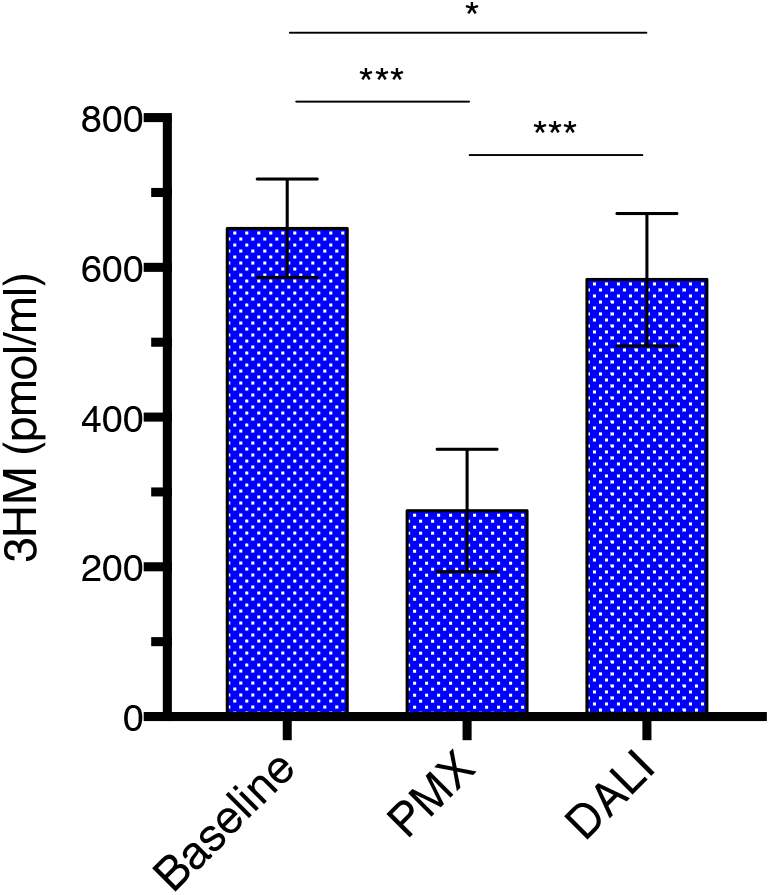
LPS levels before and after incubation of LPS-spiked, lipoprotein-deficient plasma with LDL apheresis microbeads and polymyxin microbeads. LPS levels are decreasing after incubation with polymyxin (PMX) microbeads in LPS-spiked, lipoprotein-deficient human plasma from 10 control plasma samples, as assessed by 3HM levels. LPS decrease induced by incubation with LDL apheresis microbeads (DALI) is also significant, although weak (11% versus 58% reduction with PMX). Data represent mean ± SD. *** p<.001 and ns: p>0.05 by two-tailed paired t-test.

The baseline lipid profile of plasma samples was within normal range: total cholesterol 2.05 ±0.6 g/l, triglycerides 1,24 ±0.6 g/l, HDL cholesterol 0.65 ±0.2 g/l, LDL cholesterol 1.18 ±0.5 g/l. The results obtained for LPS adsorption were not statistically significantly different between subjects with high and low levels (above and under the average) of total cholesterol, triglycerides, HDL cholesterol, LDL cholesterol. We also analyzed the relation between the percentage of LPS reduction observed with DALI beads and corresponding plasma LDL cholesterol levels, and found no statistically significant correlation (r=0.062, p=0.68). The same absence of correlation was found with triglyceride, HDL and total cholesterol levels.

### Adsorption of lipoproteins by both DALI and PMX

As expected, incubation with DALI beads led to profound alterations in lipid profile (Figure 3). LDL cholesterol (LDLc) levels were decreasing the most at only 5.1% of control, and HDL levels were not decreasing. As already known from clinical practice of LDL-apheresis (31), triglycerides levels were also markedly reduced with DALI (48.1% of control). More surprisingly, the lipid profile was also altered after incubation with PMX (Figure 3). Compared to DALI, LDLc and triglycerides reduction were more modest (55.3% and 72.7% of control, respectively), but there was also a reduction of HDL (68.5% of control). The dilution factor (based on protein levels variation from baseline) was around 0.75 for all 3 substrates (0.73, 0.77 and 0.75 for PMX, DALI and control, respectively).

**Figure 3:**
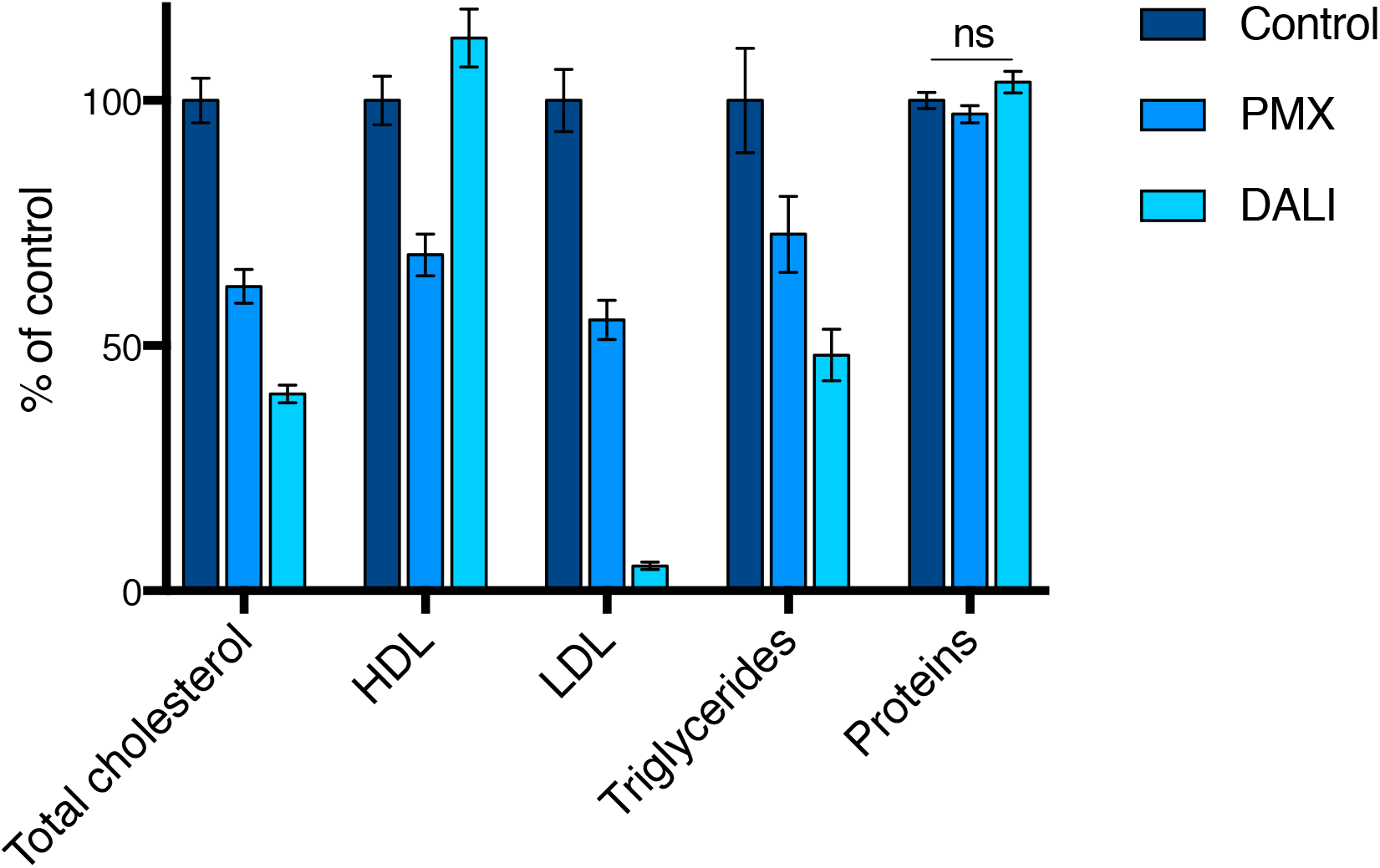
PMX beads are reducing LDL but also HDL cholesterol levels, whereas LDL apheresis beads are mostly efficient in reducing LDL and triglycerides. Data represent mean percentage reduction from baseline ± SEM. All paired comparisons for difference (3 per parameter) were statistically significant (two-tailed paired t-test p<.05), except for control and DALI® protein levels (ns: not significant).

### Efficacy of clinical LDL-apheresis to remove endogenous LPS from the circulation in patients with familial hypercholesterolemia

To determine if the effect observed in LPS-spiked human plasma could be reproduced in a clinical setting with endogenous endotoxemia, we analyzed samples from patients with familial hypercholesterolemia (FH) undergoing LDL-apheresis sessions. Blood samples were drawn before and after each session. As expected, LDLc was effectively reduced by apheresis techniques down to 26 ±9 % of baseline (Figure 4A). We analyzed samples from 77 LDL-apheresis sessions performed on 41 individual patients. Overall, LPS levels decreased between the sample drawn before and after the LDL-apheresis session, as assessed by 3HM (150.6 ±97 vs 89.7 ±70 pmol/ml, p<0.0001, Figure 4B) or LAL (0.25 ±0.15 vs 0.18 ±0.11 EU/ml, p=0.0002, Figure 4C). We compared the LPS level reduction observed with the different apheresis techniques. Most patients were treated with the Liposorber® system (Kaneka, Japan), using a column filled with microbeads coated with dextran sulfate providing negative charges to attract the apolipoprotein B present at the LDL surface (32). When looking only at the 53 (69%) Liposorber® sessions, 3HM reduction was still significant (150 ±89 vs 100.5 ±81 pmol/ml, p<0.001, Figure 4D). The second more frequently used apheresis system was the DALI® system (Fresenius, Germany). This system is based on the same basic principle of electrostatic adsorption, but uses polyacrylate instead of dextran sulfate (27). In the 19 (25%) DALI® sessions we analyzed, 3HM levels were also reduced (156.8 ±127 vs 68.4 ±37 pmol/ml, p<0.001 Figure 4E). The remaining 5 sessions (6.5%) used double-filtration plasmapheresis (DFPP). DFPP takes advantage of the large size of lipoproteins to separate them from other plasma constituents, combining a plasma filter (2 000kDa molecular weight cutoff membrane separating plasma from blood cells) with a second membrane with lower cutoff (1 000kDa) (33). This method is used in patients in whom other systems are not well tolerated or insufficient. In DFPP sessions, 3HM levels were also decreased (131.25 vs 63.4 ±12 pmol/ml, p=0.009, Figure 4F). Even though few DFPP sessions were analyzed, this result is suggesting that LPS reduction induced by LDL apheresis is due to the removal of lipoproteins themselves and not to a direct fixation of LPS by beads, e.g. by electrostatic interactions, although this can’t be ruled out by our results.

**Figure 4:**
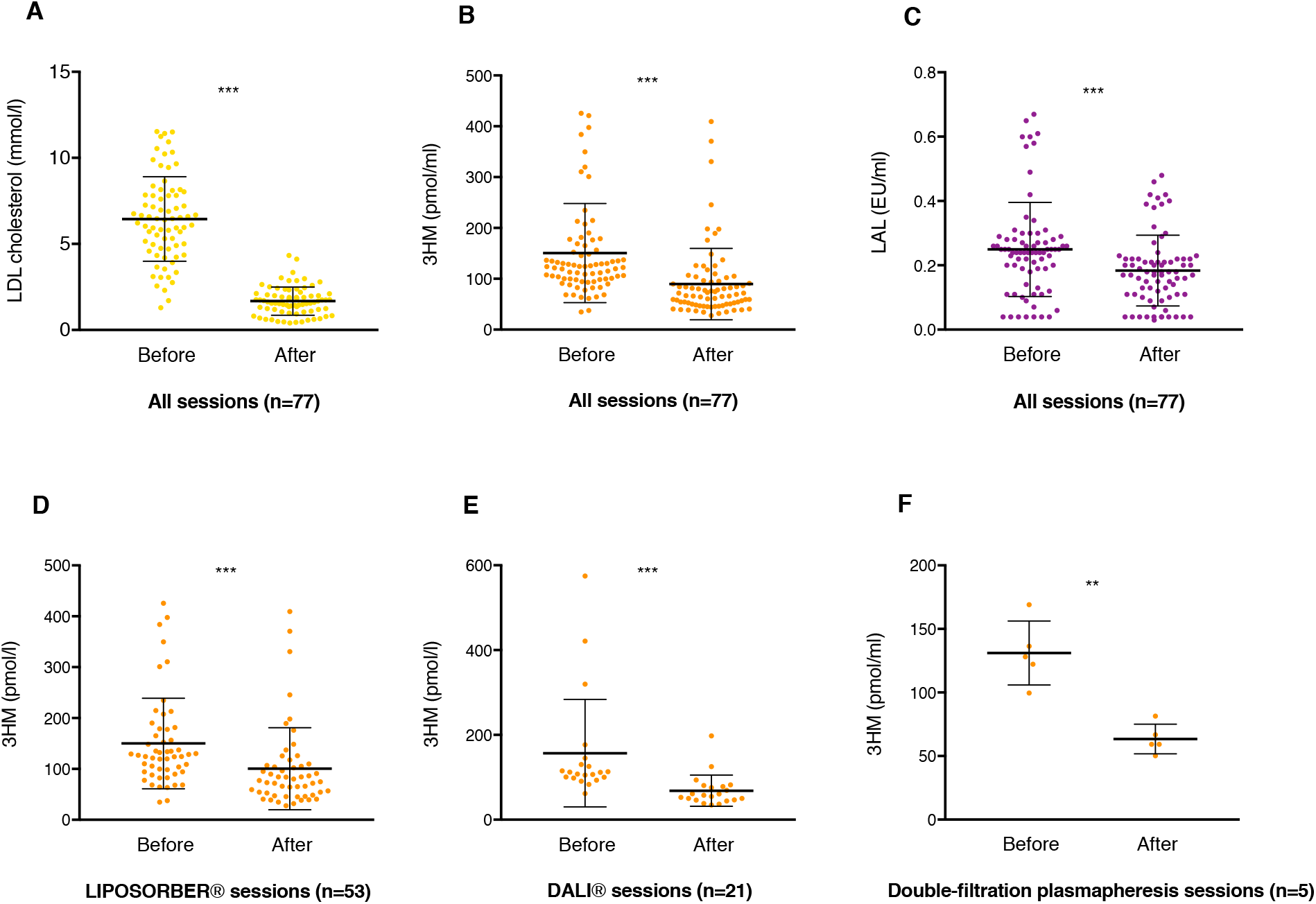
LPS levels are decreased after LDL apheresis sessions in patients with familial hypercholesterolemia. We confirmed the efficacy of these LDL apheresis sessions on LDL cholesterol levels (85,2% reduction) (A). LPS levels were decreasing after 77 LDL apheresis sessions performed in 41 patients, as assessed by 3HM levels (B) and LAL (limulus amoebocyte assay) (C). LPS reduction is significant regardless of the apheresis technique: with LIPOSORBER® (D), DALI® (E) or double-filtration plasmapheresis (F). Data represent mean ± SD. *** p<.001 and ** p<.01 by two-tailed paired t-test.

### Removal of endogenous LPS in patients presenting severe gram-negative bacteria infections

After demonstrating that LDL adsorption is efficiently removing spiked LPS from healthy human plasma, and that endogenous LPS from FH patients is also removed by LDL apheresis sessions, we sought to determine if endogenous, “pathogen-related” LPS can also be removed from the circulation. To that end, we used plasma from a cohort of critically ill patients recruited in our center. Plasma samples drawn at their admission to the intensive care unit were available for 507 patients, 60 of them with positive blood cultures, of whom 24 were positive for GNB. Finally, plasma samples (500µl aliquots stored at −80°C) from 22 patients were used (2 consent forms missing). Nineteen of them (86%) presented with septic shock criteria, the 3 other patients had severe sepsis according to sepsis-2 definitions applying at the time of study (34). The average lipid profile of plasma samples was close to normal but with high variations between patients: total cholesterol 2.51 ±1.3 g/l, triglycerides 1,76 ±0.9 g/l, HDL cholesterol 0.58 ±0.4 g/l, LDL cholesterol 1.20 ±1.2 g/l. Each of these samples was then incubated as described before for 2 hours at 37°C with DALI® microbeads (200mg of beads for 400µl of plasma), without additional LPS spiking. The small volume of available plasma per patient prevented the use of the agarose control substrate. To make up for the absence of control, the 3HM levels after incubation were corrected for each patient using measured protein dilution factor, averaging 0.83 ±0.16. Of note, the protein dilution factor was not significantly different when compared head-to-head with control substrate in previous *in vitro* experiments (Figure 3). The following correction formula was applied to each incubated sample: [corrected 3HM] = [measured 3HM] / [individual dilution factor]. Corrected 3HM levels were significantly decreased by 24% (p<.0001, Figure 5) after incubation with DALI® microbeads. This is suggesting that LDL apheresis can also remove LPS circulating during GNB-induced septic shock and sepsis.

**Figure 5:**
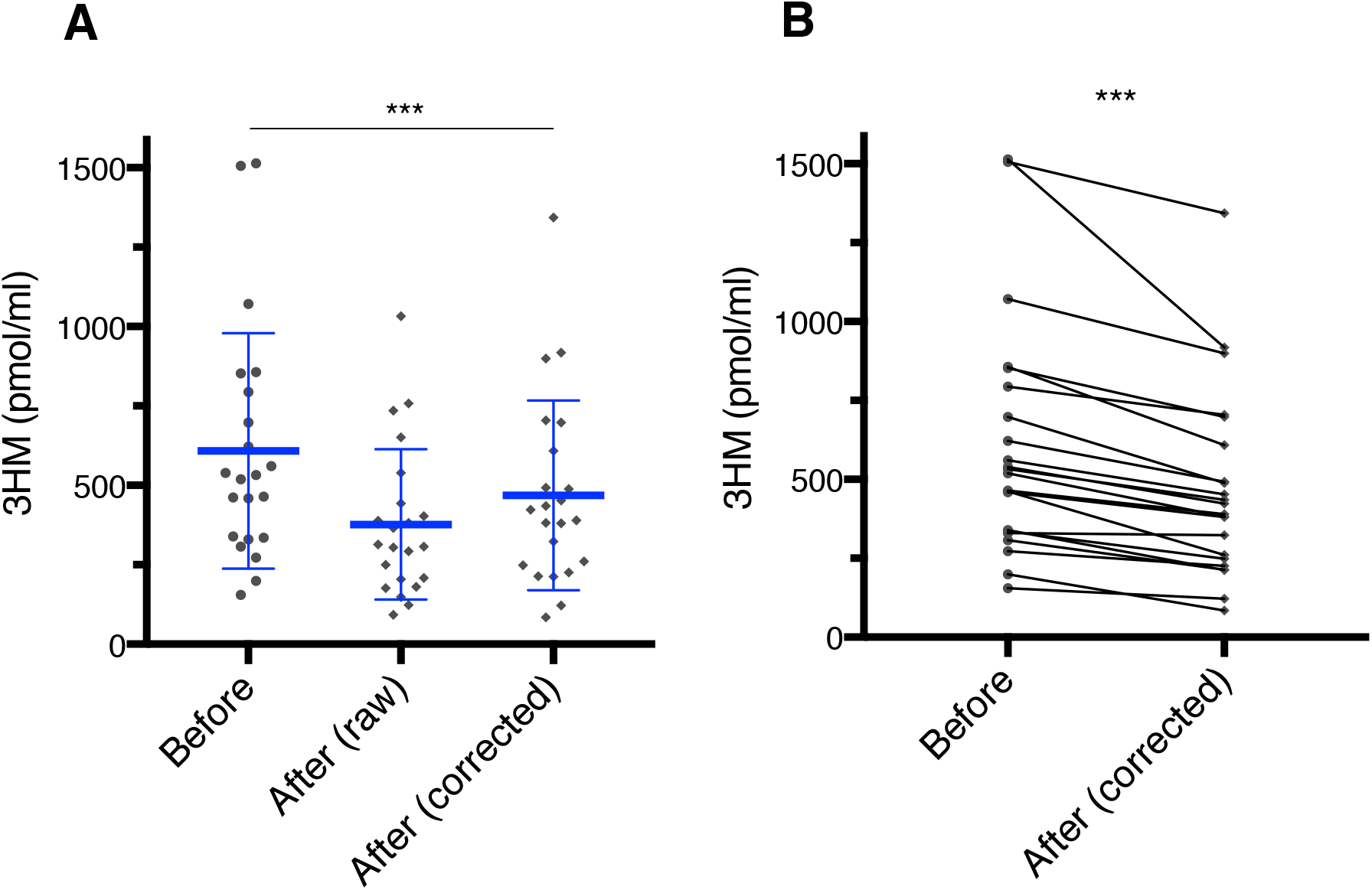
LPS reduction by LDL apheresis in the plasma of septic patients with GNB bacteremia. 3HM levels are decreased in the plasma of 22 septic patients with GNB bacteremia, even after correction of the 3HM levels after incubation with protein dilution (A). Before-after comparison shows a significant 24.1% reduction (B). *** p<.001 with two-tailed paired t-test.

Like in healthy volunteers, LPS adsorption was similar between subjects with high and low levels (above and under the average) of total cholesterol, triglycerides, HDL cholesterol, LDL cholesterol. We did not find either any statistically significant correlation between LDL, HDL, triglyceride and total cholesterol levels and the percentage of LPS reduction.

## Discussion

We demonstrated for the first time that LDL apheresis is a viable alternative to PMX adsorption for the removal of LPS from plasma. Extracorporeal LPS removal during sepsis is still in the focus of research with many recent and ongoing trials, despite disappointing results obtained with PMX hemoperfusion. LPS levels reduction with PMX hemoperfusion was low or even absent when it was measured in clinical trials (7). Our results show that LDL adsorption is almost as efficient as PMX to reduce LPS concentration in plasma spiked with a high concentration of LPS. Conversely, LPS adsorption is impaired in lipoprotein-deficient plasma, and absent in PBS. This is indicating that the LPS reduction with LDL apheresis microbeads is indeed mediated by LDL adsorption rather than by direct binding of LPS by the beads. However, although the efficiency of DALI microbeads was considerably reduced in lipoprotein-deficient plasma, 3HM was still significantly reduced. This may suggest that DALI beads can also bind “free” LPS, or LPS bound to proteins (such as LBP or PLTP). Moreover, we found that LPS reduction by DALI microbeads was not correlated to LDL levels, which may suggest that the quantity of lipoproteins is not the primary determinant of LDL apheresis efficiency to remove LPS from plasma. Thus, other qualitative aspects such as lipid and protein composition (35), as well as potential LPS “preload” of lipoproteins should be considered (10). Another question is raised by the discrepancy that we observed in vitro between the drastic reduction of LDL levels (96% ±0.3) and the relatively modest reduction of LPS levels with DALI (27% ±11%), although in vivo (in FH patients) reduction levels of LDL and LPS were closer (85% vs 40%, respectively). Thus, the LPS that is not removed with LDL may be free in plasma but also bound to other lipoproteins such as HDL, as suggested by recent data (36).

We found that PMX could also non-specifically adsorb lipoproteins, an underrecognized fact that could account for a part of its efficacy in plasma. To our knowledge, the effect of PMX adsorption on global lipoprotein levels reduction is described here for the first time. However, this effect was suggested by a study demonstrating a binding of PMX with radiomarked LDL (37). In this study, hydrophobic interaction between PMX and LDL particles could explain the observed reduction. PMX is indeed an amphiphilic molecule, with a polar (cationic) as well as a non-polar moiety. Hydrophobic bonds are also critical in the interaction between LPS and PMX (38). The hydrophobic adsorption of lipoproteins by PMX would have 2 consequences: first, it would be a new, underrecognized mode of action of PMX for reducing LPS burden (by adsorbing both the lipoprotein and the LPS molecules bound to the lipoprotein); secondly, it might competitively inhibit PMX’s direct binding of LPS in its other forms: bound to monocytes, comprised in intact bacteria or cell wall debris, micelles (1)… Then, it could also paradoxically compromise the efficacy of “free” LPS clearance from the circulation by competitive inhibition, and partly explain the poor results obtained with PMX hemoperfusion for septic shock (6, 7).

The efficacy of LDL apheresis to remove LPS was confirmed in a cohort of patients undergoing therapeutic LDL apheresis for FH. Endotoxemia is commonly associated with atherosclerosis and hypercholesterolemia (39), and LPS removal could contribute to the reduction in atherosclerosis observed in these patients (40). Lastly, LDL adsorption reduced LPS levels in plasma from septic patients with GNB bacteremia. However, as we did not directly test LDL apheresis in septic patients, our study does not allow to conclude that the LPS reduction obtained with LDL apheresis would improve clinical outcome or if adverse effects would prevail. Furthermore, it is hard to say whether LDL apheresis would even reduce *in vivo* inflammation accordingly, i.e. the removed LPS may already be out of reach of the TLR4 pathway. One should also consider the possibility that lowering plasma lipoproteins could lead to a decrease in LPS binding capacity (inhibiting “reverse LPS transport”), and subsequently increase free LPS concentration and macrophage activation. Strategies that remove free LPS may be more effective than removing LPS bound to lipoproteins, which may already be neutralized. Nevertheless, the equilibrium between free and lipoprotein-bound LPS is complex, and both could be reduced by LDL-apheresis. Another possible limitation for LDL apheresis is the fact that large amounts of LPS can also be bound to VLDL during sepsis (41, 42), which could mitigate the effect of LDL-apheresis in this context. Also, we did not find a greater effect of DALI compared to PMX, as predicted by our initial hypothesis. We cannot exclude that in a clinical setting (hemoperfusion with limited contact time with the substrate, etc.), the difference would be more pronounced in favor of one or the other. However, it is difficult to conclude, based on our results, that LDL-apheresis during sepsis would fare better than PMX hemoperfusion. Removal of LDL particles may still have an advantage over conventional PMX hemoperfusion, as lipoproteins carry many other lipidic pathogen associated molecules such as lipoteichoic acid (11).

The comparative efficacy of LDL-apheresis should be tested first in animal models of septic shock. However, extracorporeal circulation therapy is technically very difficult to apply to murine models. Overall, these results still confirm the major role of the reverse LPS transport by circulating lipoproteins, making them a prime target to study and treat endotoxemia-related conditions.

## Data Availability

Data are available upon reasonable request to corresponding author

## Abbreviations

3HM: 3-hydroxy myristate
DFPP: double-filtration plasmapheresis
FH: familial hypercholesterolemia
GNB: gram-negative bacteria
ICU: intensive care unit
LAL: limulus amoebocytes lysate
LPS: lipopolysaccharide
LTA: lipoteichoic acid
PAMPs: pathogen-associated molecular patterns
PCSK9: proprotein convertase subtilisin/kexin 9
PLTP: phospholipid transfer protein
PMX: polymyxin
PMX-DHP: polymyxin direct hemoperfusion

## Data availability statement

The data that support the findings of this study are available from the corresponding author, upon reasonable request: Dr Auguste Dargent, Médecine Intensive Réanimation, Hôpital Edouard Herriot, 69003 Lyon.

## Acknowledgements

The authors thank Pr Christiane Mousson of Dijon University hospital, for her help in obtaining DALI® cartridges. The authors thank Pr Marc Bardou, Agnès Maurer and Dr Maxime Luu, at the Centre d’Investigation Clinique Plurithématique (INSERM, CIC-P 1432), for the recruitment of healthy volunteers. The authors also thank Fiona Ecarnot (EA3920, University Hospital Besancon, France) for translation and editorial assistance. This work was supported by a French Government grant managed by the French National Research Agency under the program “Investissements d’Avenir” with reference ANR-11-LABX-0021 (LipSTIC Labex). The cohort study was funded by a French ministry of health (Programme Hospitalier de Recherche Clinique regional) grant.

